# Assessment of a Laboratory-Based SARS-CoV-2 Antibody Test Among Hemodialysis Patients: A Quality Improvement Initiative

**DOI:** 10.1101/2020.08.03.20163642

**Authors:** Dena E. Cohen, Gilbert Marlowe, Gabriel Contreras, Marie Ann Sosa, Jair Munoz Mendoza, Oliver Lenz, Zain Mithani, Pura Margarita Teixeiro, Nery Queija, Araceli Moneda, Jean S. Jeanty, Katie Swanzy, Misha Palecek, Mahesh Krishnan, Jeffrey Giullian, Steven M. Brunelli

**Affiliations:** DaVita Clinical Research, Minneapolis, Minnesota, USA; Department of Nephrology, University of Miami, Florida, USA; DaVita Inc, Denver, Colorado, USA

**Keywords:** SARS-CoV-2, COVID-19, antibody testing, hemodialysis

## Abstract

**Introduction:** The coronavirus disease 2019 (COVID-19) pandemic is caused by severe acute respiratory syndrome coronavirus 2 (SARS-CoV-2) infection. Although tests to detect anti-SARS-CoV-2 antibodies have been developed, their sensitivity and specificity in hemodialysis patients have not been previously assessed.

**Methods:** As part of a quality improvement (QI) initiative, nasopharyngeal swabs and predialysis blood samples were collected on the same day from adult patients receiving routine hemodialysis care at clinics managed by a large dialysis organization in the greater Miami, Florida region (23 – 30 Apr 2020). Polymerase chain reaction (PCR) tests for SARS-CoV-2 and chemiluminescence immunoassays for anti-SARS-CoV2 antibodies were performed according to manufacturer-specified protocols.

**Results:** Of 715 participants in the QI initiative, 38 had symptomatology consistent with COVID-19 prior to or during the initiative. Among these, COVID-19 was PCR-confirmed in 14 and ruled out in 20, with the remaining 4 being inconclusive. Among the 34 patients with known COVID-19 status, the sensitivity and specificity of the antibody test were 57.1% and 85.0% when either antibody was considered. The remaining 677 patients had no record of symptoms consistent with COVID-19, nor any known exposure. Of these, 38 patients (5.6%) tested positive for anti-SARS-CoV-2 antibodies.

**Conclusions:** The operational characteristics of the laboratory-based antibody test make it sufficient to rule in, but not rule out, SARS-CoV-2 infection in the appropriate clinical circumstance. A substantial proportion of dialysis patients may have had asymptomatic SARS-CoV-2 infection.

## Introduction

The current coronavirus disease 2019 (COVID-19) pandemic is caused by severe acute respiratory syndrome coronavirus 2 (SARS-CoV-2) infection.^1^ This pandemic may pose a particular threat to patients with end-stage kidney disease (ESKD), including those treated with in-center hemodialysis (ICHD). Such patients, by definition, cannot self-isolate; they need to attend thrice-weekly treatments at a dialysis center, to which they are often transported by a shared-ride vehicle or public transit. Because of these factors, rapid identification of ICHD patients infected with SARS-CoV-2 is an essential component of epidemic management in this population, enabling their isolation from uninfected patients during treatment.^2^

The role of antibody testing in the identification of dialysis patients with current or previous SARS-CoV-2 infection is not known. The antibody response to SARS-CoV-2 in the general population has been the subject of intense investigation, leading to an understanding of the kinetics of appearance of immunoglobulin (IgM and IgG) antibodies in terms of time since symptom onset.^3^ On average, IgM antibodies appear first, often within a week of symptom onset, and diminish over time such that by 5-6 weeks after symptom onset, most patients have reverted to an IgM negative state.^4-6^ IgG antibodies, by contrast, tend to appear somewhat later, often 1-2 weeks after symptom onset, but are sustained for an extended period thereafter;^4-6^ given the recent nature of the COVID-19 pandemic, the absolute duration of IgG positivity cannot yet be known.

The extent to which findings from the general population with respect to antibody levels and kinetics can be extrapolated to patients with ESKD is unclear, given the functionally immunocompromised state of such patients.^7^ Thus, although numerous tests to detect IgM and IgG antibodies against specific SARS-CoV-2 proteins have received Emergency Use Authorization from the Food and Drug Administration (FDA), the potential utility of such tests among patients treated with ICHD has not yet been clarified.

Here, we report the findings of a quality improvement (QI) initiative conducted with the goal of assessing the operational feasibility of utilizing a laboratory-based SARS-CoV-2 antibody test among patients treated with ICHD. The sensitivity and specificity of the antibody test were then assessed in the subset of participating patients with known status with respect to SARS-CoV-2 infection. The frequency of antibody positivity was determined among patients who had neither a history of symptoms consistent with COVID-19, nor known exposure to the virus.

## Methods

### Patients

Eligible patients were adults (≥18 years of age) receiving dialysis care at a large dialysis organization operating in the greater Miami, Florida, region (23-30 April 2020). Patients with and without a recorded history of symptoms consistent with COVID-19 were included. Patients in the former category were classified as either having COVID-19 or having the diagnosis of COVID-19 excluded, based on the totality of available clinical evidence and polymerase chain reaction (PCR) test results (referred to throughout as COVID-19^+^ and COVID-19^-^, respectively). Patients in the latter category had no recorded history of either symptoms consistent with COVID-19, nor known contact with a person infected with SARS-CoV-2.

All study data were derived from statistically deidentified electronic medical records. Therefore, according to title 45, part 46 of the US Department of Health and Human Services’ Code of Federal Regulations, this study was deemed exempt from institutional review board (IRB) or ethics committee approval (Quorum IRB, Seattle, WA). We adhered to the Declaration of Helsinki and informed consent was not required.

### SARS-CoV-2 PCR and Antibody Testing

As part of the QI initiative, samples for reverse transcription quantitative PCR and antibody analysis were collected on the same day for each patient. PCR samples were collected using sterile flocked nasopharyngeal swabs that were stored refrigerated in viral transport medium prior to shipment. PCR tests were performed using 2 primer and probe sets (Fulgent Genetics). Tests were scored as positive if both SARS-CoV-2 sequences were detected, negative if neither sequence was detected, and inconclusive if only 1 region was detected.

Blood samples for antibody analysis were collected prior to dialysis treatment in a 5-mL serum separation tube, clotted for 30 minutes, centrifuged, and refrigerated prior to shipment. Indirect chemiluminescence immunoassays for anti-SARS-CoV2 IgM and IgG antibodies (Diazyme Laboratories, Inc) were performed according to the manufacturer’s protocol at a centralized, accredited clinical laboratory (DaVita Labs). The tests detect antibodies against the SARS-CoV-2 spike and nucleocapsid proteins. Per the manufacturer’s recommendation, samples were scored as IgM positive or IgG positive if the corresponding test reading was >1 arbitrary unit (AU)/mL, and negative otherwise.

### Analysis

Patient characteristics were summarized as of the sample collection date. COVID-19 status was ascertained among the subset of patients in whom universal clinic entrance screening had detected symptoms consistent with COVID-19. Such patients subsequently underwent diagnostic workup per clinical protocol, which included at least one nasal PCR test. Those patients who ruled in for COVID-19 by PCR testing were ascribed as known COVID-19 positive (COVID-19^+^); those patients who eventually ruled out for COVID-19 by serial PCR testing were ascribed as known COVID negative (COVID-19^-^). Among patients with known COVID-19 status, sensitivity (true positives/COVID-19^+^ patients) and specificity (true negatives/COVID-19^-^ patients) were calculated overall; to account for potential biological latency in antibody production, a sensitivity analysis was conducted that considered only those antibody tests conducted 3 or more days after symptom onset. Antibody levels (expressed as AU/mL) were plotted relative to the number of days that elapsed between recorded date of symptom onset and sample collection.

Seroprevalence of SARS-CoV-2 infection was assessed in a separate group of patients who had never screened positive on universal clinical entrance screening for symptoms or high-risk contacts. Among these patients, counts of antibody-positive patients were summarized, categorized on the basis of the PCR test result obtained using the swab collected on the same day as the serum sample. Confidence intervals (CIs) were estimated using the binomial (exact) method where indicated.

## Results

A total of 715 patients participated in the QI initiative (Figure 1). Of these, 38 had a history of symptoms consistent with COVID-19: 14 were COVID-19^+^ and 20 were COVID-19^-^. Status with respect to COVID-19 could not be unambiguously determined in the remaining 4 patients, who were thus excluded from the subsequent analysis. The remaining 677 patients had no record of symptoms consistent with SARS-CoV-2 infection or any known exposure. Participant characteristics are summarized in Table 1.

**Table 1.**
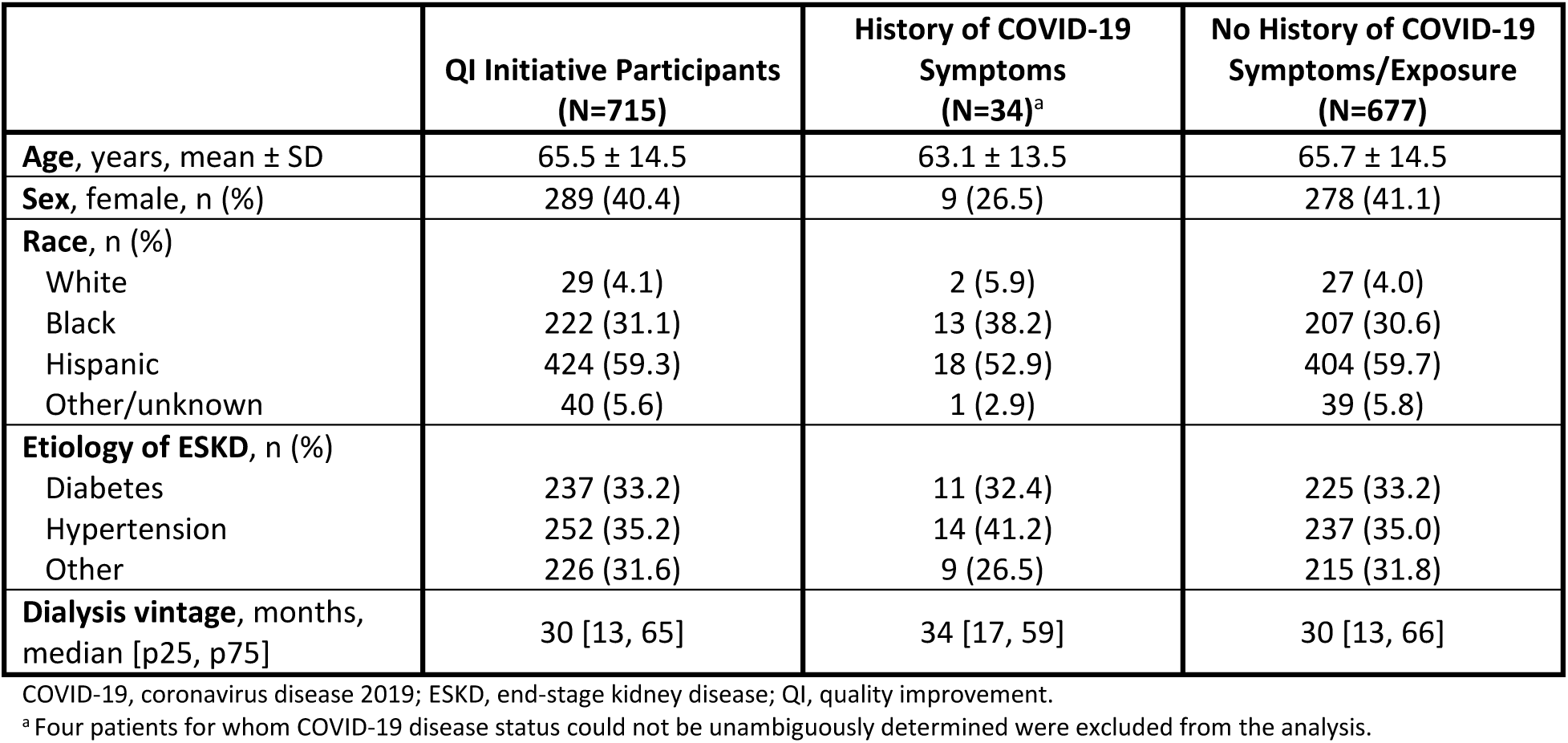
Characteristics of patients who participated in the quality improvement initiative

|  | QI Initiative Participants<br>(N=715) | History of COVID-19<br>Symptoms<br>(N=34)<sup>a</sup> | No History of COVID-19<br>Symptoms/Exposure<br>(N=677) |
| --- | --- | --- | --- |
| **Age**, years, mean ± SD | 65.5 ± 14.5 | 63.1 ± 13.5 | 65.7 ± 14.5 |
| **Sex**, female, n (%) | 289 (40.4) | 9 (26.5) | 278 (41.1) |
| **Race**, n (%) |  |  |  |
| White | 29 (4.1) | 2 (5.9) | 27 (4.0) |
| Black | 222 (31.1) | 13 (38.2) | 207 (30.6) |
| Hispanic | 424 (59.3) | 18 (52.9) | 404 (59.7) |
| Other/unknown | 40 (5.6) | 1 (2.9) | 39 (5.8) |
| **Etiology of ESKD**, n (%) |  |  |  |
| Diabetes | 237 (33.2) | 11 (32.4) | 225 (33.2) |
| Hypertension | 252 (35.2) | 14 (41.2) | 237 (35.0) |
| Other | 226 (31.6) | 9 (26.5) | 215 (31.8) |
| **Dialysis vintage**, months,<br>median [p25, p75] | 30 [13, 65] | 34 [17, 59] | 30 [13, 66] |

COVID-19, coronavirus disease 2019; ESKD, end-stage kidney disease; QI, quality improvement.
<sup>a</sup> Four patients for whom COVID-19 disease status could not be unambiguously determined were excluded from the analysis.

**Figure 1.**
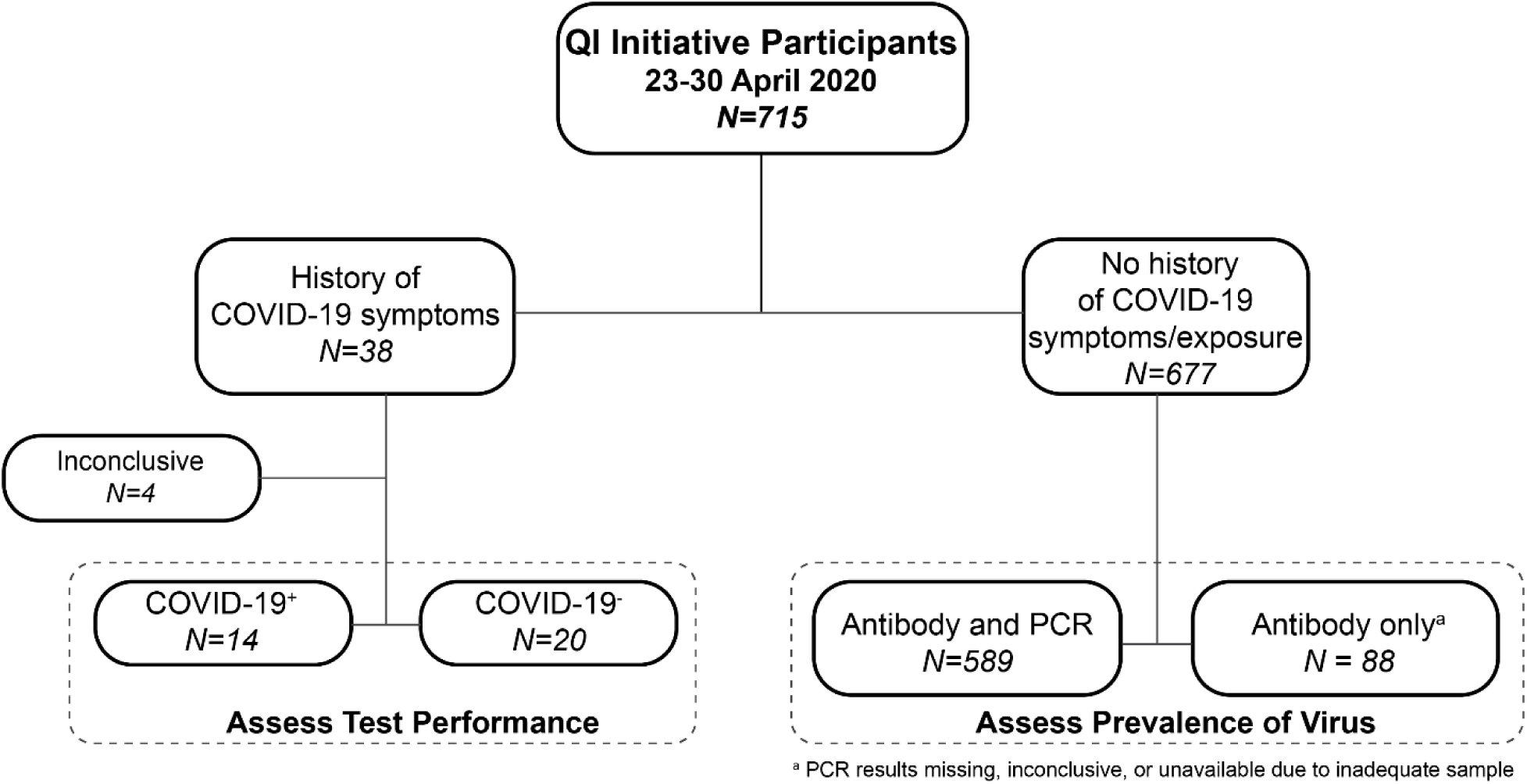
QI initiative cohort. A diagram of the disposition of participants in the QI initiative (N=715) is shown. Performance of the antibody test was assessed in participants with known COVID-19 status. Prevalence of SARS-CoV-2 infection was assessed among participants with no history of COVID-19 symptoms or exposure. COVID-19 indicates coronavirus disease 2019; PCR, polymerase chain reaction; QI, quality improvement; SARS-CoV-2, severe acute respiratory syndrome coronavirus 2.

### Assessment of test performance in patients with history of symptoms consistent with COVID-19

To assess the sensitivity and specificity of the antibody test, results were analyzed among the 14 COVID-19^+^ and 20 COVID-19^-^ participants. When a positive test result was considered as the presence of either an IgM or an IgG value over the manufacturer-specified threshold of 1 AU/mL, the sensitivity of the test was 57.1%, with specificity of 85.0% (Table 2). Considering IgM alone and IgG individually, sensitivities were 35.7% and 53.9%, respectively; specificities were 95% and 85%, respectively. These calculations were repeated on the subset of serum samples taken 3 or more days following symptom onset, yielding sensitivity and specificity of 66.7% and 84.2%, respectively. Considering IgM and IgG individually, sensitivities were 33.3% and 66.7%, respectively; specificities were 94.7% and 84.2%, respectively. Levels of IgM and IgG antibodies were examined as a function of the number of days since symptom onset (Figure 2). Although numbers are sparse, no participants tested positive for IgM beyond 38 days after symptom onset. Unexpectedly, among 8 COVID-19^+^ participants who underwent serum collection on the date of symptom onset, 2 tested positive for both IgM and IgG and 1 additional participant tested positive for IgM only.

**Table 2.**
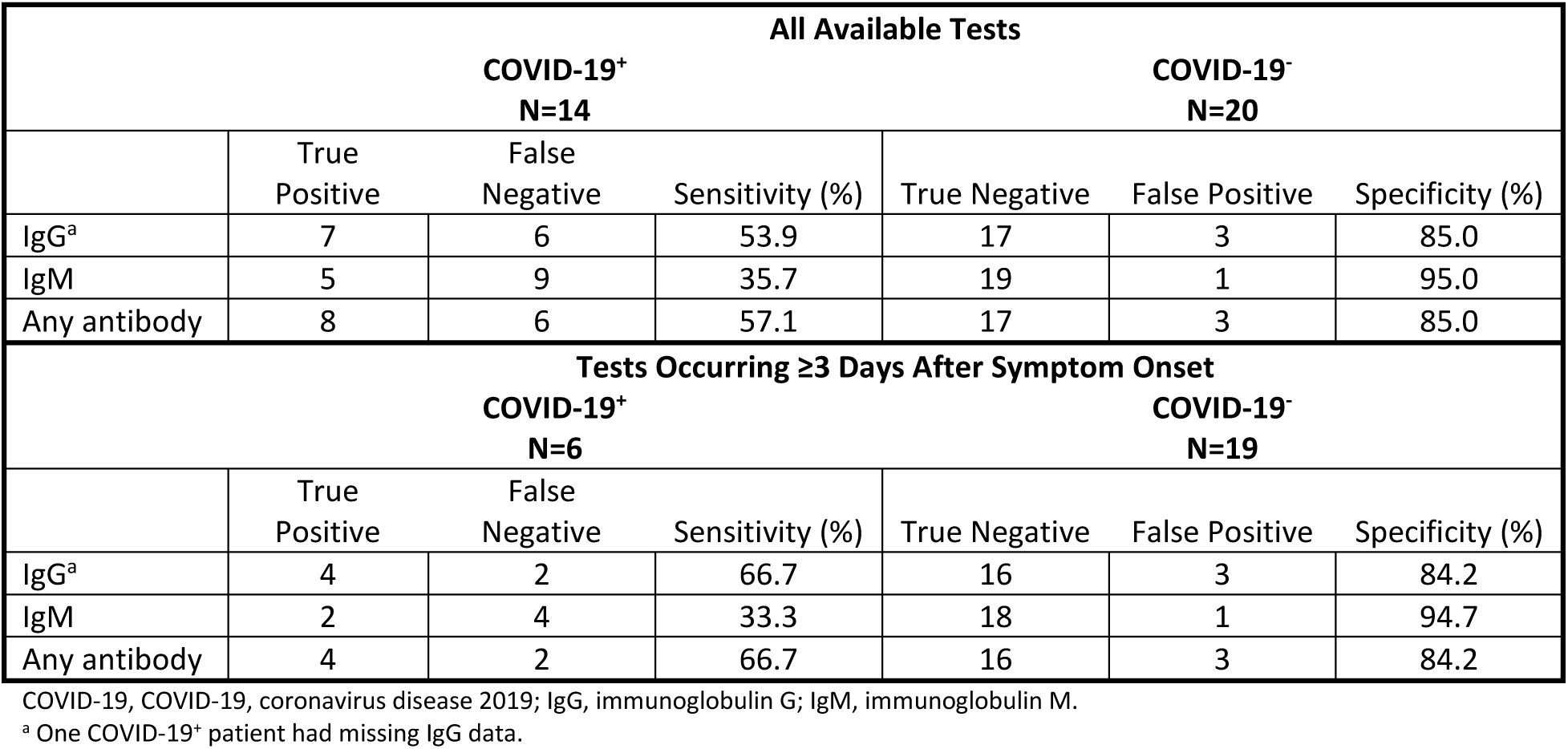
Sensitivity and specificity of antibody test among patients with known COVID-19 status

**Figure 2.**
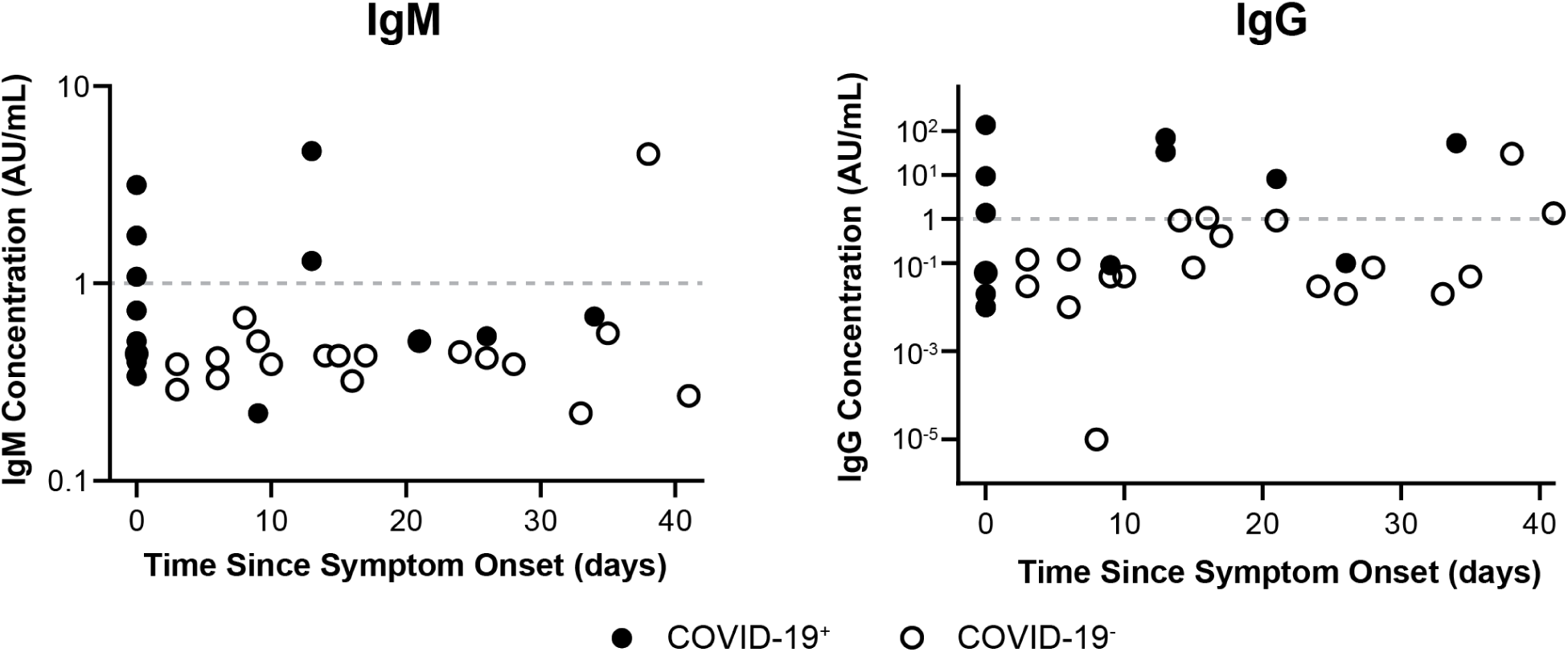
IgG and IgM concentrations among QI initiative participants. Measured IgM (left panel) and IgG (right panel) concentrations among COVID-19^+^ (filled circles) patients and COVID-19^-^ (open circles) patients are shown on the basis of elapsed time in days between symptom onset and serum sample collection. The manufacturer-specified cut-off between positive and negative test results was 1 AU/mL (dashed grey line). AU indicates arbitrary units; COVID-19, coronavirus disease 2019; IgG, immunoglobulin G; IgM, immunoglobulin M; QI, quality improvement

### Assessment of SARS-CoV-2 prevalence in patients without history of symptoms consistent with COVID-19

Among 677 participants in the QI initiative who had no record of symptoms consistent with COVID-19 or known viral exposure, 588 had a negative PCR test for SARS-CoV-2, 88 had unavailable PCR data, and 1 had a positive PCR result (Table 3). Based on measured IgG and IgM levels, 38 patients were antibody positive and 639 were antibody negative, corresponding to a seropositivity rate of 5.6% (95% CI 4.0-7.6%) among patients with neither symptomatology nor PCR evidence of SARS-CoV-2 infection. Taking both antibody and PCR testing into account, 5.8% (95% CI 4.1-7.8%) of patients in the QI initiative with no history of COVID-19 symptoms or exposure had biological evidence of current or prior SARS-CoV-2 infection.

**Table 3.**
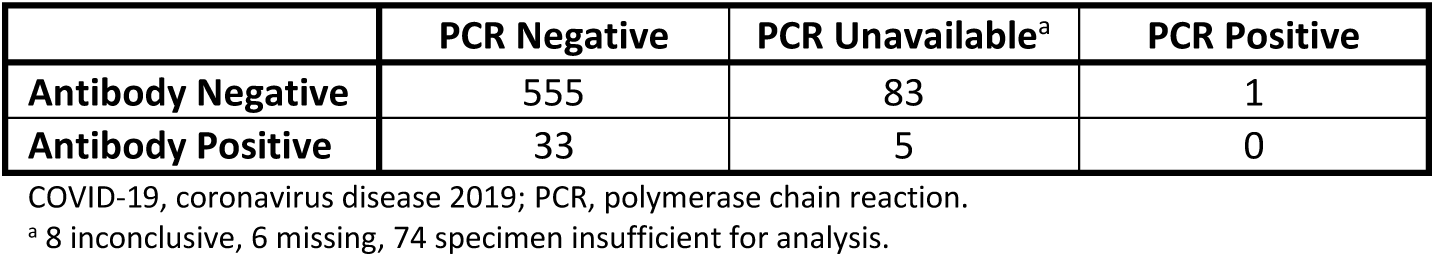
PCR and antibody test results among patients with no history of COVID-19 symptoms or exposure

|  | PCR Negative | PCR Unavailable<sup>a</sup> | PCR Positive |
| --- | --- | --- | --- |
| Antibody Negative | 555 | 83 | 1 |
| Antibody Positive | 33 | 5 | 0 |

COVID-19, coronavirus disease 2019; PCR, polymerase chain reaction.
<sup>a</sup> 8 inconclusive, 6 missing, 74 specimen insufficient for analysis.

## Discussion

A QI initiative was undertaken at a large dialysis organization to assess the operational feasibility of a laboratory-based assay for anti-SARS-CoV-2 antibodies. This initiative demonstrated that collection and shipment of samples for antibody testing during routine dialysis care is straightforward and that the test can be readily conducted at scale at a clinical laboratory. Antibody test results were available as soon as or more rapidly than PCR results, opening the possibility of simultaneously considering both test types to evaluate patient status with respect to SARS-CoV-2 infection.

The data generated by the QI initiative also provided an opportunity to examine the accuracy of the antibody assay and assess its potential utility in guiding patient care decisions. The high operational specificity of the antibody assay, 85.0%, supports its use to supplement PCR testing to rule in the disease in the assessment of symptomatic patients of unknown COVID-19 status. Specifically, among newly symptomatic patients, the use of positive serologic test results as a criterion for isolating patients as COVID-19^+^ could be considered. Although the current study cannot directly address the question of whether the assay cross-reacts with antibodies against other coronaviruses, the high specificity observed among COVID-19^-^ patients argues that such cross-reactivity is limited.

It is important to note that the low sensitivity of the antibody test, measured here as 57.1%, represents the functional sensitivity of the assay among the COVID-19^+^ patients in the QI initiative and is not an intrinsic characteristic of the assay per se. Not all ESKD patients seem to mount an antibody response to SARS-CoV-2, so negative antibody testing may reflect immunologic anergy rather than assay failure. Similar anergy in the ESKD population has been observed in other circumstances, such as in response to hepatitis B vaccination.^8^ Additionally, the kinetics of the immune response may render patients antibody negative at the time of testing, despite SARS-CoV-2 infection. Although we attempted to address one aspect of this in a sensitivity analysis, considering only patients 3 or more days post symptom onset, this approach does not address heterogeneity among patients with respect to antibody kinetics. Likewise, these analyses did not directly consider waning IgM levels at later times and probably underestimate the sensitivity of IgM in a biologically appropriate time window. Further studies, in which COVID-19^+^ ICHD patients are sampled repeatedly over a period of weeks or months following symptom onset, are needed to further clarify these issues. Regardless, given the functional sensitivity observed here, negative serologic testing should not be considered as evidence of the absence of disease, and serologically negative patients should undergo diagnostic workup as otherwise indicated.

Finally, the QI initiative provided an opportunity to assess the burden of asymptomatic SARS-CoV-2 infection among ICHD patients who had no record of either symptoms or exposure to the virus. The seroprevalence among such participants (5.6%, 95% CI 4.0-7.6%) was quite similar to what was reported in the general population of Miami, Florida, contemporaneously (6.0%, 95% CI 4.4-7.9%).^9^ This suggests that, despite the fact that ICHD patients are not able to adopt social distancing practices to the same degree as the general population, this did not substantially increase the burden of SARS-CoV-2 infection. Both data points indicate that the rate of biological infection with SARS-CoV-2 is substantially greater than the known rate of COVID-19 disease, which was reported to be approximately 0.01% at that time,^10^ underscoring the need for universal precautions.

Together, these data support use of a laboratory-based antibody test to supplement PCR testing of ICHD patients for evaluation of status with respect to SARS-CoV-2 infection. This study should be interpreted in the context of its limitations. The number of COVID-19^+^ and COVID-19^-^ patients available for analysis was small. Patients were sampled at only one time point, precluding longitudinal analysis of antibody concentrations following symptom onset. The date of symptom onset among symptomatic patients was abstracted from data collected for administrative purposes. The results reported here pertain only to the specific antibody test used and cannot be generalized to other laboratory-based assays or to point-of-care lateral flow immunoassays.

## Data Availability

Underlying study data are proprietary to DaVita, Inc

## Author contributions

Study design: KS, MK, JG, SMB. Clinical leadership and execution: GC, MAS, JMM, OL, ZM, PMT, NQ, AM, JSJ. Data acquisition: KS, GM. Data analysis: DEC, SMB. Preparation of manuscript: DEC, SMB. All authors reviewed and provided critical feedback on the manuscript and agree to be accountable for all aspects of the work.

## Acknowledgments

We wish to acknowledge all members of the Outcomes Research group at DaVita Clinical Research for helpful conversations. We also acknowledge the key contributions of all clinical and laboratory staff members who made this work possible. Finally, we acknowledge the patients who participated in this initiative, without whom the work would not have been possible.

